# Automated Detection of Sleep Apnea Using Machine Learning: A Novel Approach Using Smartphone and Microphone for Breathing Sound Analysis

**DOI:** 10.1101/2024.03.22.24304619

**Authors:** Om Bhatt

## Abstract

In this study, we evaluate the accuracy of a novel setup in the detection of apneas and hypopneas and estimating the apnea-hypopnea index (AHI). The study device setup consists of a microphone placed underneath the nose and a smartphone to collect the data. We recruited patients who were referred to the St. Josephs Hamilton Sleep Clinic for a sleep study. Data from our study device is collected simultaneously with polysomnography (PSG) in the sleep lab. A total of 26 patients were recruited, of which 2 dropped out during the data collection. Data from the microphone was too noisy for interpretation in 3 patients. Across the remaining 21 patients, the AHI based on their PSG ranged from 2 to 125 events/h, with an average AHI of 34 events/h. We used regression models trained on microphone audio data to identify noise and we developed an algorithm based on root-mean-square of the audio data for automatic detection of apneas and hypopneas. With reference to the PSG, our study device had a sensitivity of 94% and specificity of 87% in detecting apneas/hypopneas across a cumulative 120.9 hours of sleep data and more than 3700 such events. Our study device was able to accurately predict the AHI to within 4.3 events/h (+/-3.1). As such, we can conclude that our study device is accurate in identifying apneas/hypopneas, estimating the AHI and at ruling out cases of severe sleep apnea and can potentially be used as a screening test for this purpose. Our study device has the practical advantages of being very low cost and potentially more accessible as a screening tool, though further validation studies are needed to study accuracy across a larger population and for use at-home.

## Background / rationale

Obstructive sleep apnea is a highly prevalent condition (roughly 15% of all men, 5% of all women)^i,ii^ with a high symptom burden as well as high associated cardiovascular risk. Patients suspected of having OSA are referred for a sleep study, or polysomnography (PSG), for assessment. PSG is a comprehensive but expensive test to administer. We would like to investigate whether breathing sounds recorded using a microphone placed under the nose can be used to detect variations in breathing patterns. This “study device”, consisting of a microphone and a smartphone for recording and storing breathing sounds, would represent a low-cost and more accessible alternative if it found to be accurate. This device can be used by patients in their own home using their personal smartphone. Previous studies have demonstrated that patients who have home sleep apnea assessment are more likely to adhere to treatment and have better control of their symptoms and comorbidities^iii,iv^.

## Research question

How accurate are measurements of breathing sounds recorded using a microphone placed under the nose in detecting clinically significant apneas and hypopneas as compared to a polysomnography?

## Study design

The objective of this early phase study is to collect data using the study device alongside simultaneous PSG in any patient who is referred for a sleep study without exclusion. We compare the accuracy of our study device in detecting apneas and hypopneas as identified by the PSG data interpreter. This also allows us to compare the accuracy of the study device in diagnosing sleep apnea with the gold standard and report on the sensitivity and specificity for various outcomes.

Our study proposition was reviewed and approved by the Hamilton Integrated Research Ethics Board.

## Data collection

Patients over age 18 years who had been referred to the clinic for suspected sleep apnea were approached for participation in the study. STOP-BANG scores were assessed for all participants who provided informed signed consent to participate.

Patients were set up for their PSG in the usual fashion. Next, our study device was attached to the patient, as shown in Figure 1, by the sleep technician. Patients were given the option of wearing the smartphone on their arm or their waist.

**Figure 1.**
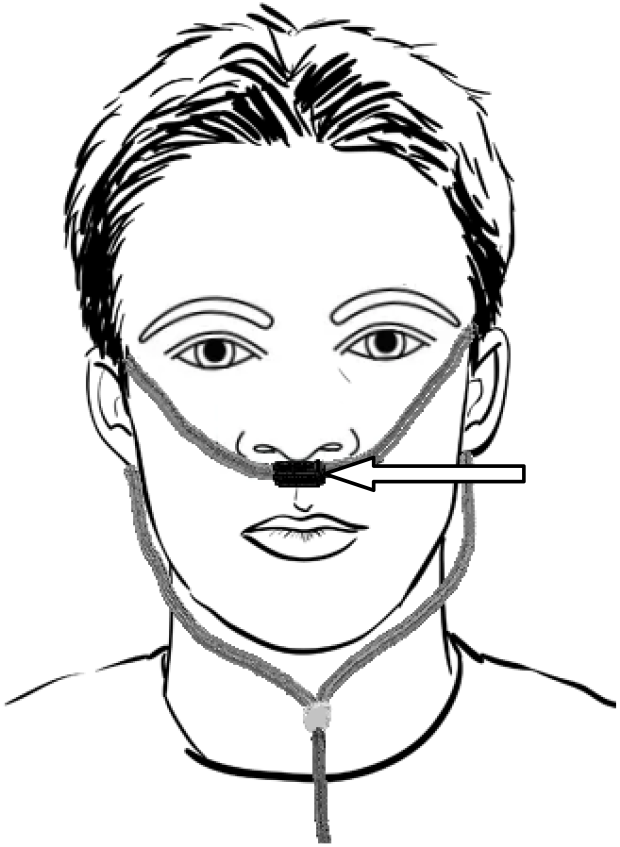
Image illustrating how the microphone (indicated by the white arrow) was placed on participants.

Our study device data included 3 axis accelerometer data, recorded at 20 Hz using the “Physics Toolbox” smartphone app which is freely available on the Google Playstore. Monophasic audio was collected from the microphone at 11025 Hz using the “Voice Recorder” app, also freely available on the Google Playstore.

## Recruitment

A total of 26 patients were recruited for this study, between the ages of 20 and 78 years, of which 18 were male. Median age was 45 years.

Two participants withdrew from their sleep study and hence also dropped out of the study.

PSG data quality was adequate for analysis for the remaining 24 patients. The PSG data was analyzed in the usual fashion by sleep technicians.

AHI was determined and categorized in the usual fashion and of the 24 patients, 10 had severe, 5 had moderate and 4 had mild AHI scores.

## Results

### Noise removal

All 24 patients had complete data collection from the study device. One patient requested to have a fan running during his sleep study and hence the audio data collected by our microphone was noisy and excluded from our analysis.

The first and last 15 minutes of the data is disregarded from analysis as this is typically when the calibration takes place and had a lot of artifactual data.

A 240Hz high pass filter is applied to the data to remove a broad low frequency noise that seems specific to the sleep clinic environment. This noise was of high amplitude but fortunately bandlimited. Figure 2 demonstrates the audio data pre- and post-filter, showing qualitatively that the signal to noise ratio improves noticeably.

**Figure 2.**
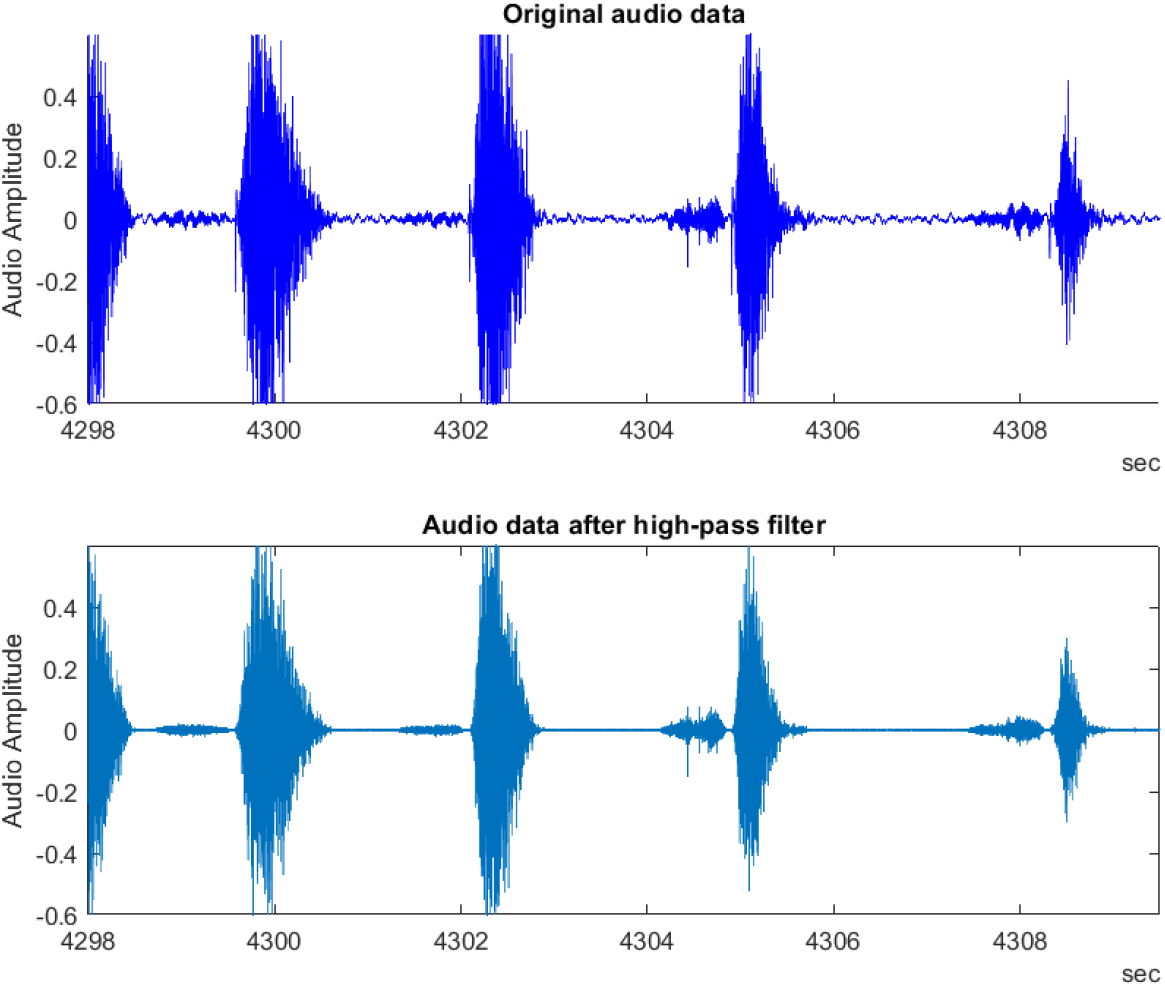
Comparison of audio data pre- and post-240 Hz high pass filter. The filter effectively removes background noise and noticeably improves the signal-to-noise ratio.

Manual qualitative assessment of the device audio data revealed 4 sessions with portions of noisy audio data consistent with the microphone cable connection with the smartphone becoming loose. An example of this is shown in Figure 3. This data was manually labelled, along with 3 hours of clean audio data. and a classification model was trained to automatically identify these periods.

**Figure 3.**
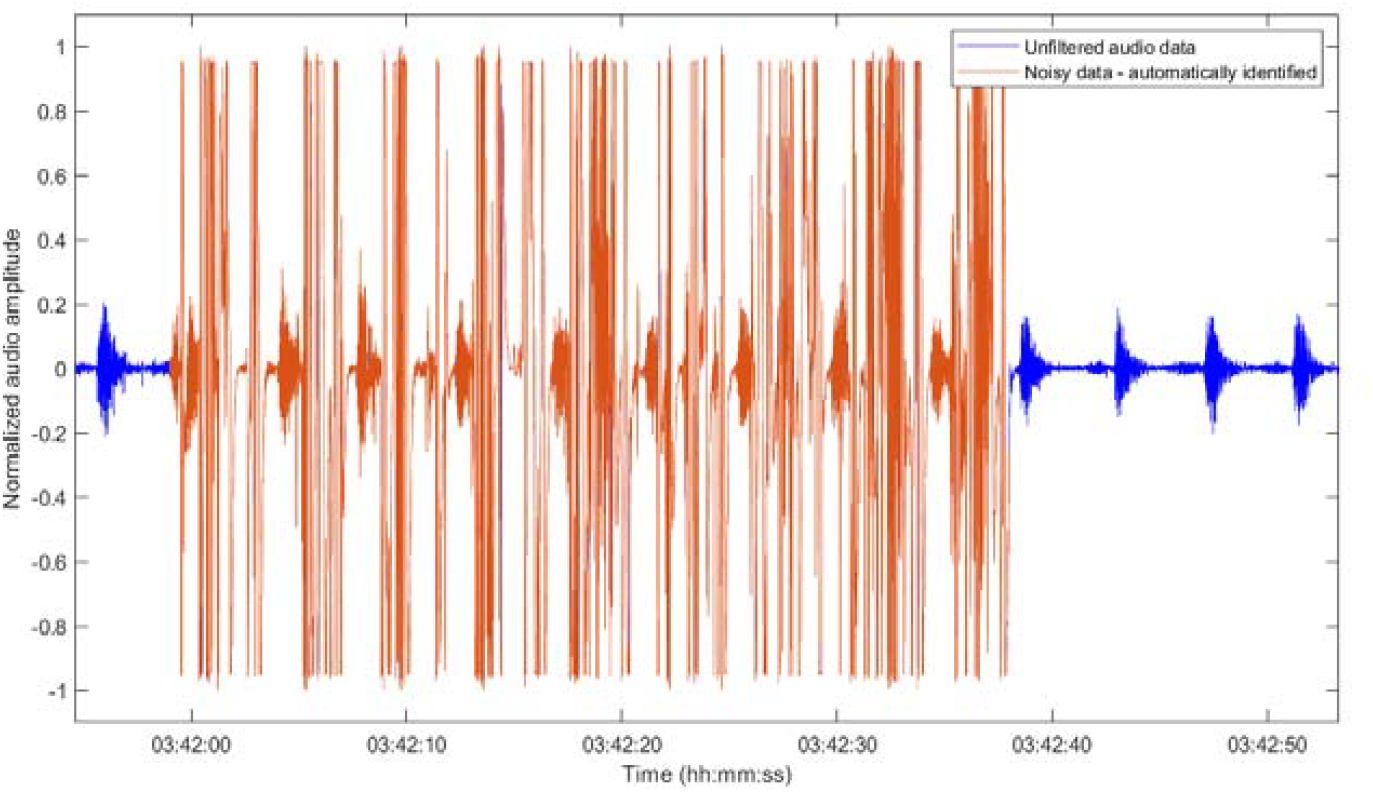
Noise in the audio data is automatically identified and noisy segments are removed prior to further analysis. In this instance, noise is due to the microphone cable coming loose from the phone. Data shown is from participant 22, where there was too much noise present in the recording and the entire session had to be excluded from analysis.

For 3 of these sessions, noise was present throughout the entire recording from the microphone. Hence, these 3 sessions were excluded from further analysis.

Finally, the RMS of the accelerometer data was computed over 2 second windows and a threshold was determined that corresponded to patient movement. The audio data during periods of movement were also excluded from analysis for identifying apneas/hypopneas.

### Apnea/hypopnea episodes detection

Audio data is split into 0.3 second overlapping windows and the RMS is calculated for these windows. Then, we search for the maximum RMS value in 1.5-second-long windows, which is the loudest bit of audio. As illustrated in Figure 4, these maximum values correspond to instances of breathing.

**Figure 4.**
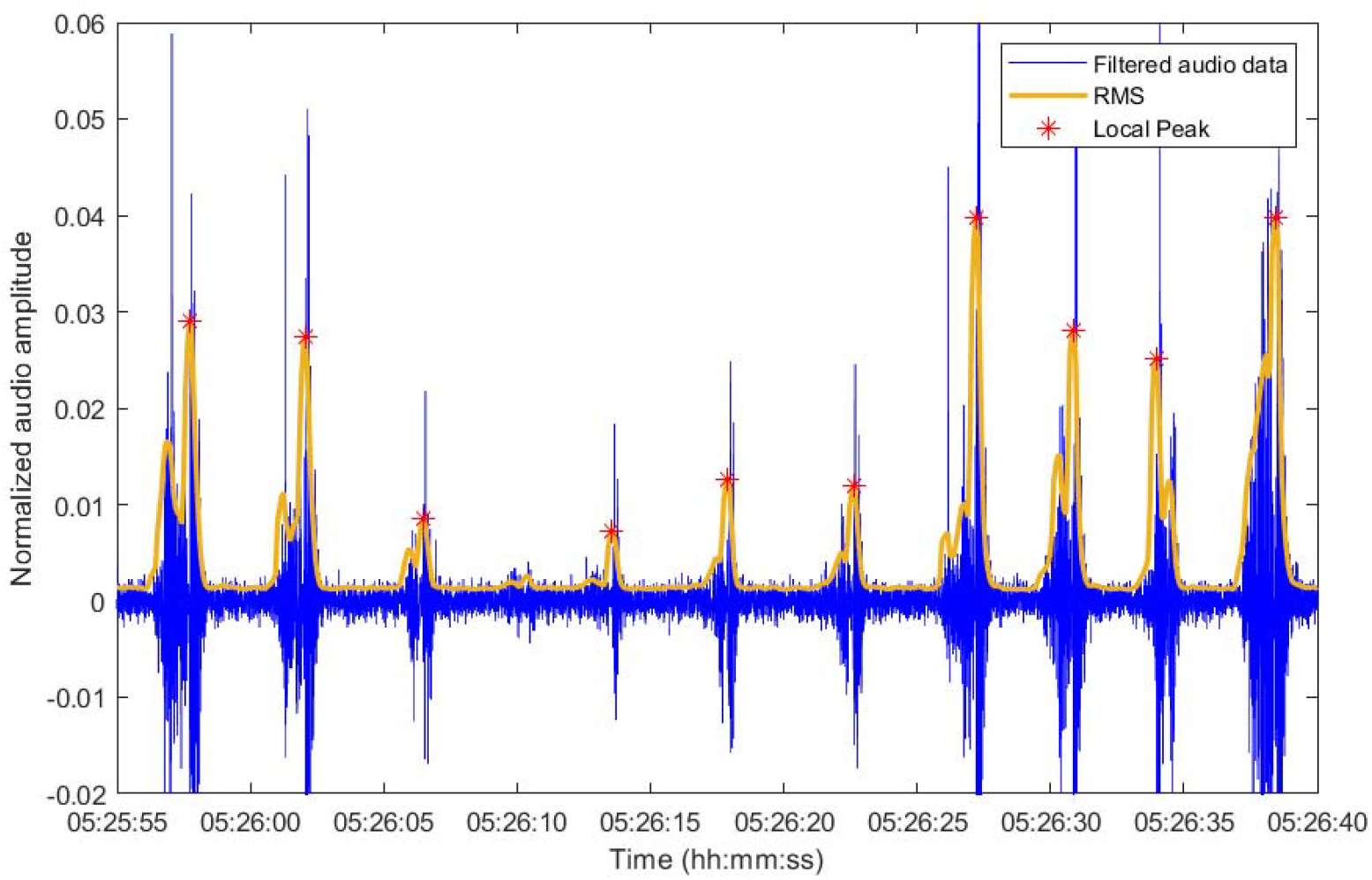
Filtered audio data and the calculated RMS, along with the peaks identified by searching for the maximum value in 1.5 second windows. The RMS and peaks are used to determine periods of quiet or absent breathing.

**Figure 5.**
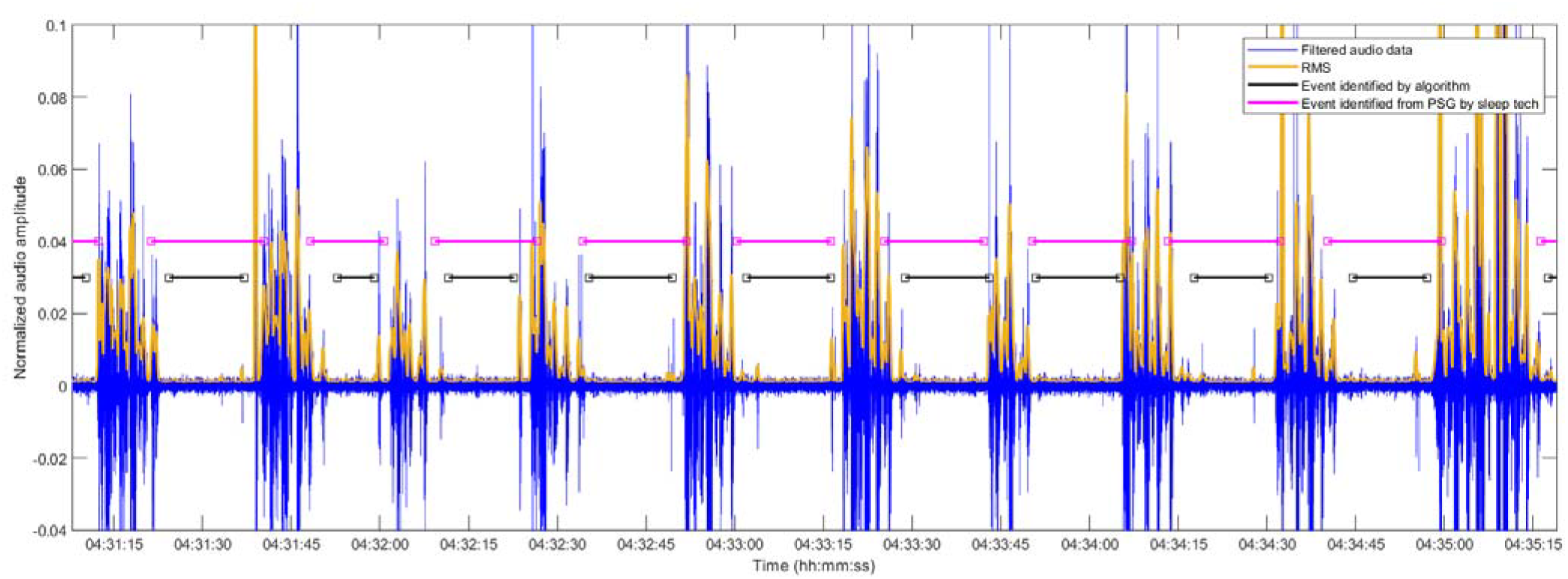
A comparison of events (apneas and hypopneas) identified by our algorithm, marked by a black line, and events identified through the PSG by a sleep technician. Qualitatively, there appears to be very good agreement between the two.

**Figure 6.**
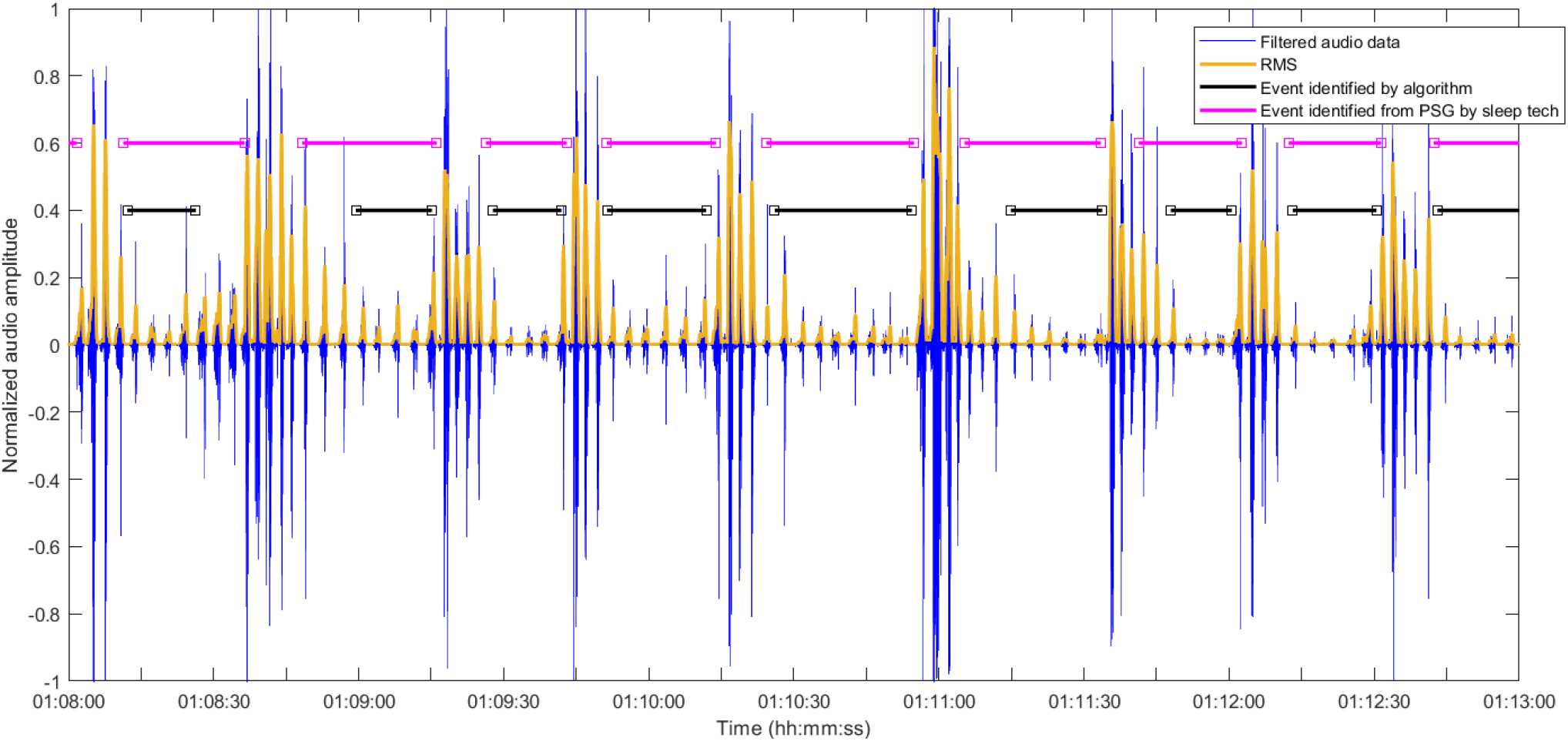
Comparison of events (hypopneas) identified by our algorithm, marked in black, and events identifed from the PSG by a sleep technician.

**Figure 7.**
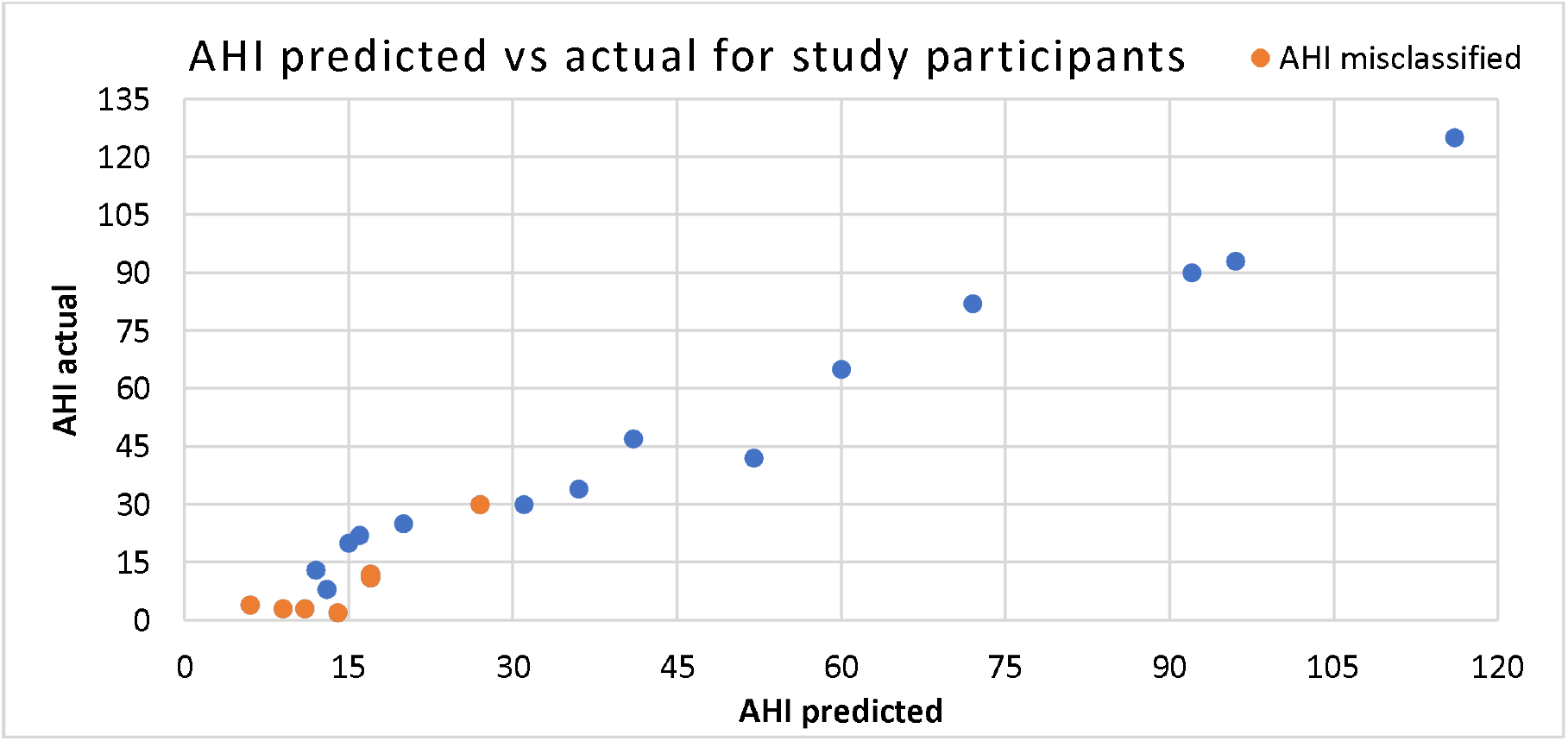
Graph comparing the AHI determined from the PSG (AHI actual, vertical axis) to the AHI predicted by our algorithm (AHI predicted, horizontal axis). Most of the misclassification errors are in participants with AHI less than 30.

**Figure 8.**
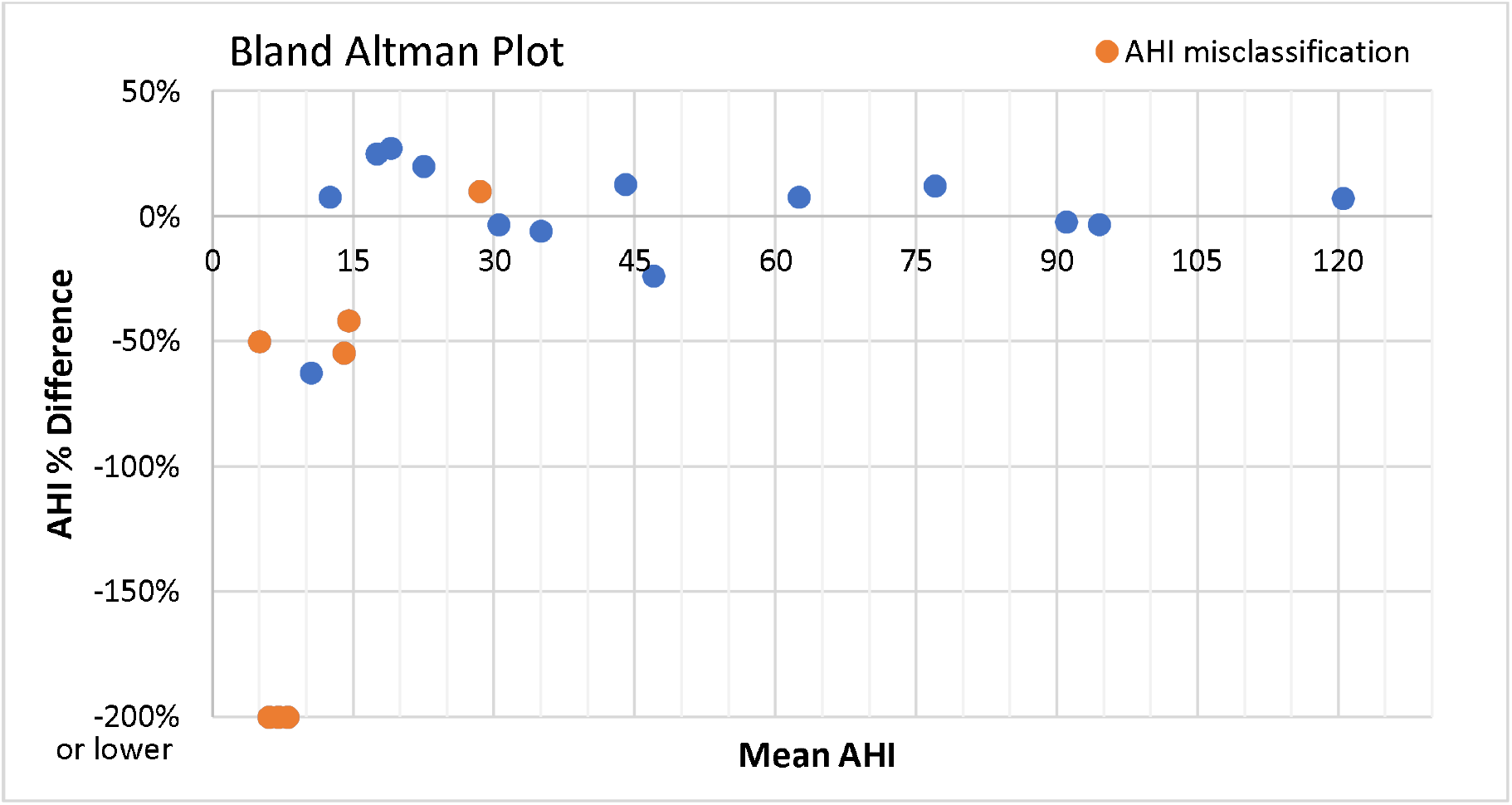
Bland Altman Plot, an alternative way of visually comparing the predicted AHI with the AHI determined from the PSG, across the 21 participants.

Once we have identified the peaks in audio data corresponding to breathing, we then compare the amplitude of the breath sound over 10 seconds to the average of the breath sounds over the previous 3 minutes. We search for periods of at least 10 seconds where the breathing volume is less than 25% of the average breathing volume in the preceding 3 minutes, and this corresponds to a apnea or hypopnea event. We also search for periods of at least 10 seconds where the breathing volume is undetectable, i.e., at the same level as background noise. The start and end times of these events are saved. We analyze all audio data and then exclude events that occur when data was noisy as noted above.

### Comparison of PSG and device event detection

Apnea and hypopnea events marked on the PSG data were extracted for each participant. Obstructive, central and mixed apneas as well as hypopneas are all combined and treated in the same way. The AHI and the total sleep time was extracted from the sleep study reports.

There is a synchronization offset between the smartphone device time and the PSG data time that is random and on the order of 2-3 seconds. Hence, our approach to see if the PSG and algorithm events match is to use the start and end times and check for any overlap. For each event identified by the algorithm, if there is a corresponding event in the PSG where the start/end times overlap, it is considered a **true positive**. If no corresponding event was noted in the PSG data, then it is considered a **false positive**.

Lastly, we split the entire audio data into 30 second segments and we label a segment as negative if there are no events identified during that segment by our algorithm. If there is an AH event in the PSG data overlapping with this segment, this segment considered a **false negative**. Else, it is considered a **true negative**.

Across the 21 patients included in the analysis, the average sensitivity and specificity of the algorithm for detecting are 91% (±9%) and 83% (±17%) respectively. Across more than 120 hours of cumulative sleep data, 3733 apnea/hypopnea events were identified from the PSG data. Our algorithm identified 3503 true positive events, giving a sensitivity of 94% in aggregate.

The PSG data is divided into epochs of 30 second duration. In total more than 14500 epochs were analyzed, of which 9893 epochs had no apnea/hypopnea events detected in the PSG data. Similarly, our study device data was divided into epochs and 8640 were true negative epochs where no events were detected by our algorithm and in the PSG data, giving a specificity of 87% across all data.

### Comparison of AHI estimate

The AHI is calculated by dividing the number of events identified from our device data by the number of hours of noise-free data analyzed. The mean absolute difference in the AHI estimate from our study device and the AHI estimate from the PSG is 4.3 events/h (±3.1).

AHI severity classification is determined with the clinically accepted thresholds of 5, 15, and 30 events/h for mild, moderate, and severe sleep apnea respectively. Across the 21 patients, the AHI severity for 7 patients was misclassified, giving a misclassification rate of 33%. Of these, 4 patients were misclassified as mild when their AHI according to the PSG was normal (less than 5); two were misclassified as moderate when their AHI according to the PSG was mild. One patient had an AHI of exactly 30 based on the PSG (i.e. severe), but a predicted AHI of 27 (i.e. moderate), and hence was technically misclassified although the difference in AHI is small.

**Table 1.** Confusion matrix comparing the algorithm predicted AHI severity compared to the AHI severity determined from the PSG, across the 21 participants.

| Predicted AHI Severity | Actual AHI severity |  |  |  |  |
| --- | --- | --- | --- | --- | --- |
|  | Total: 21 | Normal (4) | Mild (4) | Moderate (3) | Severe (10) |
|  | Normal (0) | 0 | 0 | 0 | 0 |
|  | Mild (6) | 4 | 2 | 0 | 0 |
|  | Moderate (6) | 0 | 2 | 3 | 1 |
|  | Severe (9) | 0 | 0 | 0 | 9 |

### Explorative work

Application of machine learning algorithms has the potential to greatly expand the utility of the data collected by our device. Breathing phases (inhale, exhale) can be predicted with a decision tree model trained on features from the audio data. For training the model, first the breathing phase is inferred from the nasal pressure data from the PSG (positive deflection is inhale, negative deflection is exhale). A 3-class classifier is trained, with the 3 classes being inhale, exhale and pause. The results are shown below in Figure 9. Unfortunately, the model from one patient does not generalize well to other patients.

**Figure 9.**
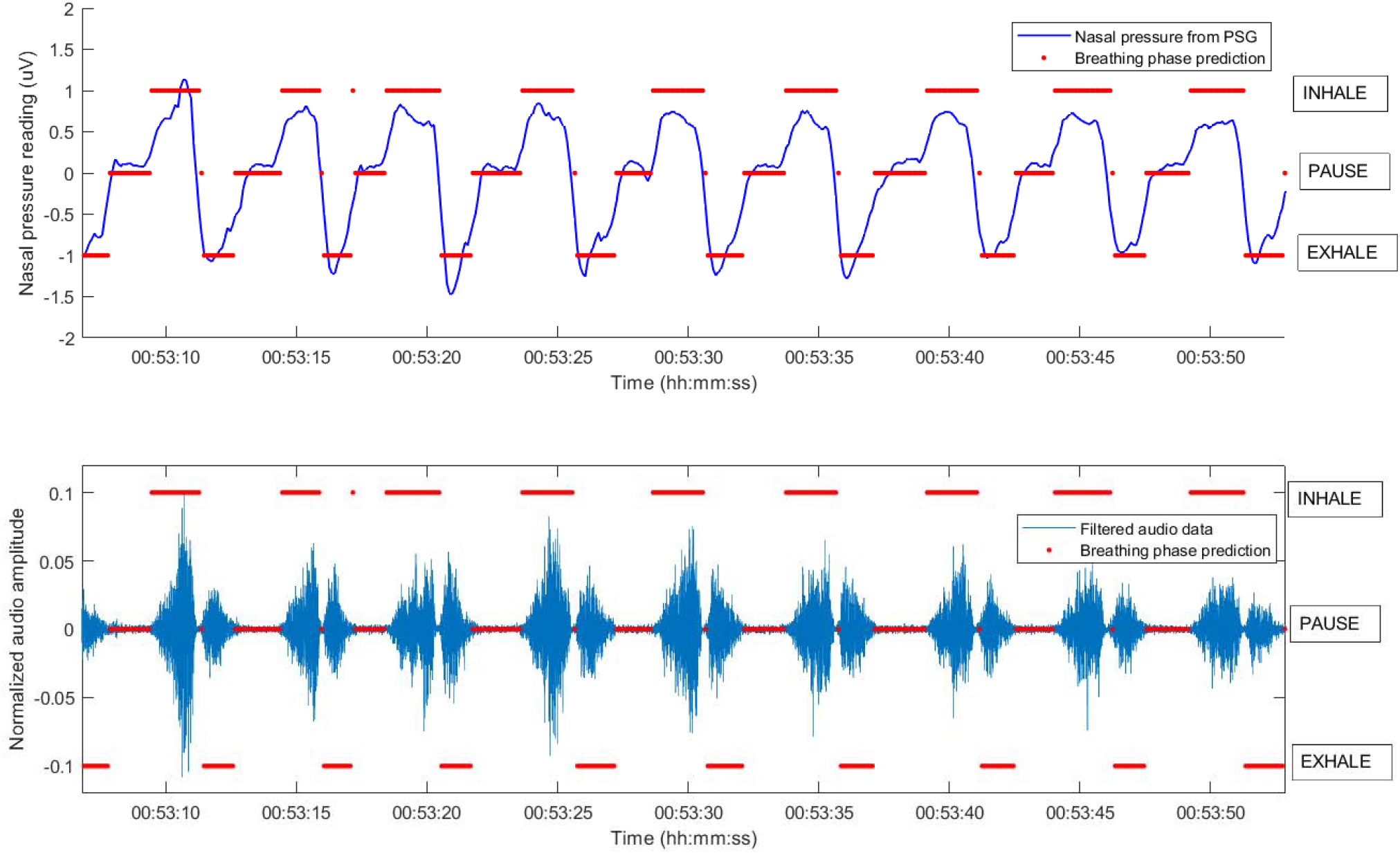
Machine learning classification algorithm, developed with features from the audio data and with the nasal pressure data as the reference, used to automatically classify data as either inhale, pause or exhale.

**Figure 10.**
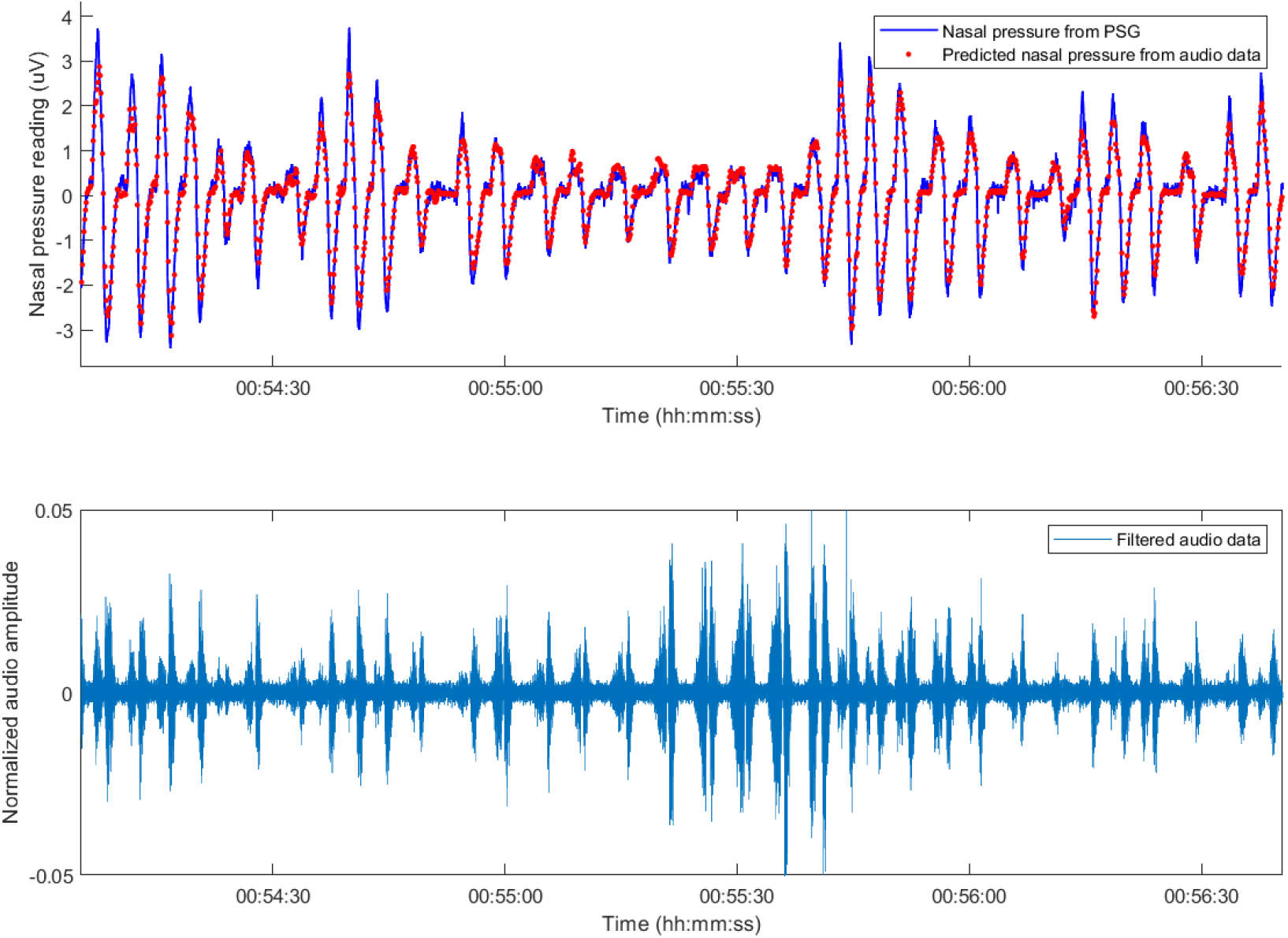
Machine learning algorithm predicts nasal pressure accurately (shown in red) from audio data (lower plot). Although the audio data between 55:00 and 55:30 shows noisy breathing, there is reduced airflow during this period, indicating likely upper airway resistance leading to stridorous noisy breathing.

Interestingly, the audio data can also be used to predict the nasal pressure directly based on a combination of features. The figure below illustrates the results of a linear regression model trained to predict the “nasal pressure” from a combination of features from the audio data collected on patient 5. Unfortunately, the trained model from a single patient did not generalize well when applied to other patients. We expect that with more sophisticated training structure and models, that a model for predicting nasal pressure tracing across all patients can be developed. The discernable advantage of this method is that periods of noisy breathing that are in fact due to upper airway obstruction (i.e. stridorous breathing) can be differentiated from breathing with appropriate airflow. This can be explored further with future studies.

## Conclusions

There were no hardware related data collection issues with our study device. With respect to data quality, 3 of 24 patients (12.5%) had poor quality data recorded. In one case, it was due to environmental noise (a fan running in the background), and in the remaining cases it was because of the microphone connection coming loose. Importantly, this type of noise is easy to identify and exclude from analysis. In comparison, less than 5% of PSG studies have issues with data quality requiring repeat assessment^v^, since typically a sleep technician can intervene during the night to rectify the problem.

Possible ways to mitigate data quality issues with our device can include a) real time signal processing in the smartphone that alerts patients to make adjustments; and b) collecting data across multiple nights. Efficacy of such strategies can be studied in a future trial.

Our algorithm for automatic detection of apneas and hypopneas from microphone data is similar to previously published algorithms based on nasal pressure tracing^vi^. Importantly, the events identified by our automated algorithm matched well with the events identified through the PSG. The high sensitivity and specificity rates suggest that our algorithm is identifying the same clinically significant event as the PSG. Certainly, a significant limitation is that the apneas and hypopneas are considered equally, and future work can include refining the algorithm to differentiate between the two to provide greater insights into the AHI score.

Across the 21 patients, our study device was able to accurately predict the AHI to within 4.3 events/h (±3.1). The AHI severity was misclassified for 7 of 21 patients. Only 1 case of severe sleep apnea was misclassified as moderate, although the difference in AHI was only 3 events/h and the AHI estimate from the PSG was exactly on the cut-off at 30 events/h. As such, we can conclude that our study device is accurate at ruling out cases of severe sleep apnea and can potentially be used as a screening test for this purpose. However, more data is needed to conclude this with full confidence.

There are number of practical advantages to our study device that motivated this work. The cost of our study device is <$30 CAD, excluding the cost of the smartphone used. In 2018, 88% of Canadians over age 15 years owned a smartphone^vii^.

Another significant advantage is ease of use, as our device can potentially be used in patient’s own home without requiring a dedicated facility or specialized staff to administer the test. Finally, patients can be evaluated over multiple nights, and this would help to reduce the number of incomplete studies and improve diagnostic accuracy. Recent studies indicate that up to 20% of patients diagnosed with a single-night study may be misclassified.

Although other at-home sleep apnea testing devices exist, they remain quite expensive, and no standalone application exists to automatically analyze the data to determine the AHI.

These practical advantages can greatly increase the accessibility and lower the cost for sleep apnea testing. Certainly, the use of this device at home by patients should be studied further in future work. Additionally, the efficacy of a standalone smartphone App that collects and analyzes the data to estimate the AHI should also be studied further in future work.

## Data Availability

All data produced in the present study are available upon reasonable request to the authors

## Appendix 1

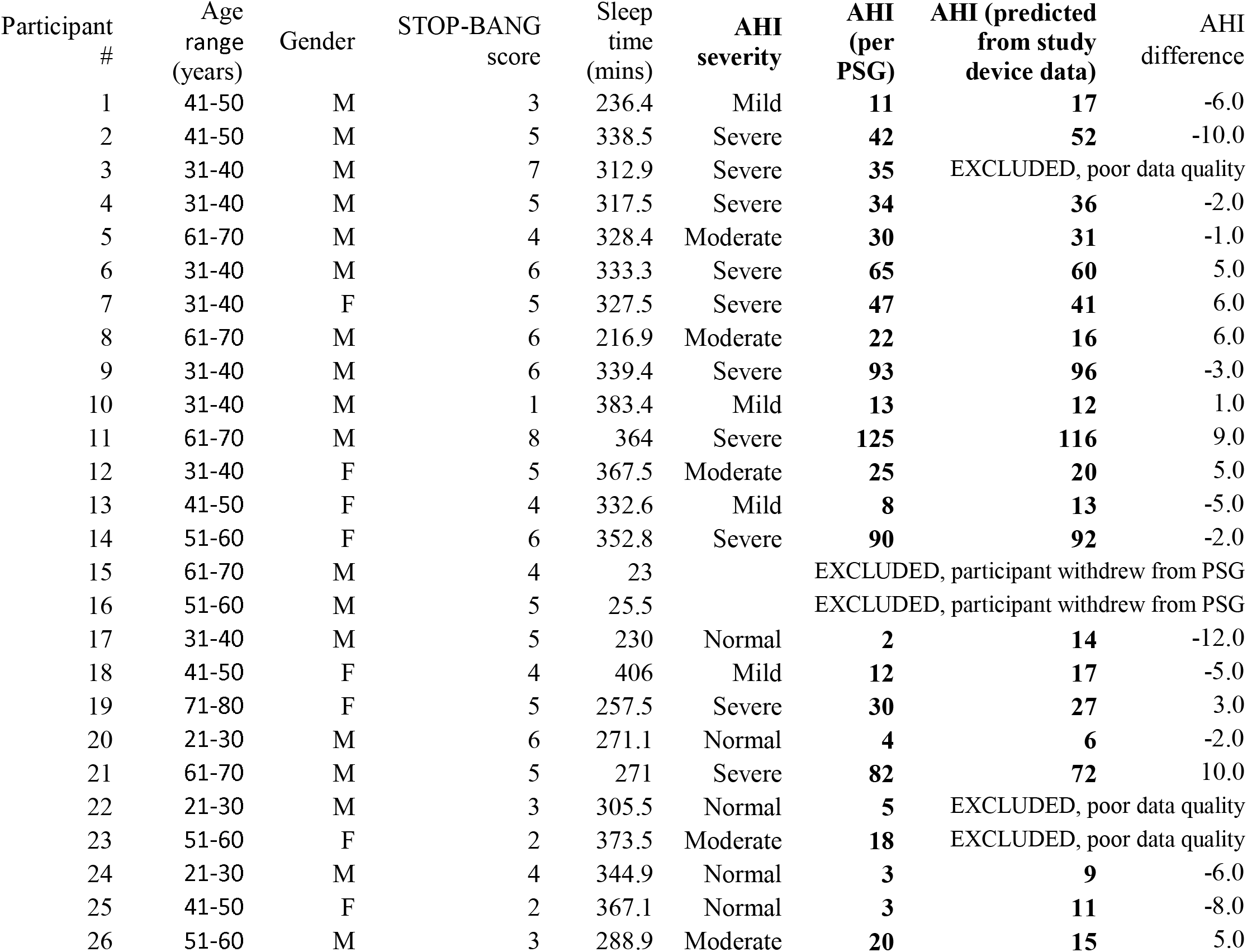

| Participant # | Age range (years) | Gender | STOP-BANG score | Sleep time (mins) | AHI severity | AHI (per PSG) | AHI (predicted from study device data) | AHI difference |
| --- | --- | --- | --- | --- | --- | --- | --- | --- |
| 1 | 41-50 | M | 3 | 236.4 | Mild | **11** | **17** | -6.0 |
| 2 | 41-50 | M | 5 | 338.5 | Severe | **42** | **52** | -10.0 |
| 3 | 31-40 | M | 7 | 312.9 | Severe | **35** | EXCLUDED, poor data quality |  |
| 4 | 31-40 | M | 5 | 317.5 | Severe | **34** | **36** | -2.0 |
| 5 | 61-70 | M | 4 | 328.4 | Moderate | **30** | **31** | -1.0 |
| 6 | 31-40 | M | 6 | 333.3 | Severe | **65** | **60** | 5.0 |
| 7 | 31-40 | F | 5 | 327.5 | Severe | **47** | **41** | 6.0 |
| 8 | 61-70 | M | 6 | 216.9 | Moderate | **22** | **16** | 6.0 |
| 9 | 31-40 | M | 6 | 339.4 | Severe | **93** | **96** | -3.0 |
| 10 | 31-40 | M | 1 | 383.4 | Mild | **13** | **12** | 1.0 |
| 11 | 61-70 | M | 8 | 364 | Severe | **125** | **116** | 9.0 |
| 12 | 31-40 | F | 5 | 367.5 | Moderate | **25** | **20** | 5.0 |
| 13 | 41-50 | F | 4 | 332.6 | Mild | **8** | **13** | -5.0 |
| 14 | 51-60 | F | 6 | 352.8 | Severe | **90** | **92** | -2.0 |
| 15 | 61-70 | M | 4 | 23 | EXCLUDED, participant withdrew from PSG |  |  |  |
| 16 | 51-60 | M | 5 | 25.5 | EXCLUDED, participant withdrew from PSG |  |  |  |
| 17 | 31-40 | M | 5 | 230 | Normal | **2** | **14** | -12.0 |
| 18 | 41-50 | F | 4 | 406 | Mild | **12** | **17** | -5.0 |
| 19 | 71-80 | F | 5 | 257.5 | Severe | **30** | **27** | 3.0 |
| 20 | 21-30 | M | 6 | 271.1 | Normal | **4** | **6** | -2.0 |
| 21 | 61-70 | M | 5 | 271 | Severe | **82** | **72** | 10.0 |
| 22 | 21-30 | M | 3 | 305.5 | Normal | **5** | EXCLUDED, poor data quality |  |
| 23 | 51-60 | F | 2 | 373.5 | Moderate | **18** | EXCLUDED, poor data quality |  |
| 24 | 21-30 | M | 4 | 344.9 | Normal | **3** | **9** | -6.0 |
| 25 | 41-50 | F | 2 | 367.1 | Normal | **3** | **11** | -8.0 |
| 26 | 51-60 | M | 3 | 288.9 | Moderate | **20** | **15** | 5.0 |

